# Mental Health Status among the South Indian Pharmacy Students during Covid-19 Pandemic’s Quarantine Period: A Cross-Sectional Study

**DOI:** 10.1101/2020.05.08.20093708

**Authors:** S Vidyadhara, A Chakravarthy, A Pramod Kumar, C Sri Harsha, R Rahul

## Abstract

**Introduction:** The COVID-19 outbreak created a major panic among all the citizens of the country owing to its severity, contagiousness within the community, lack of specific treatment and possibility of re-infection. All these factors along with the uncertain behaviour of the virus lead to state of fear and concern all throught out the nation. The current study represents the mental health survey conducted on the students of South Indiaafter the completion of one month quarantine period of the COVID-19 outbreak.

**Methodology:** The present study is a cross-sectional, web-based online survey which consists of 21-item DASS questionnaire. Thiswas used to assess the emotional states of depression, anxiety, and stress. Using Google Forms, the questionnaire was randomly distributed among the pharmacy students of selected colleges. Mean with standard deviation was calculated for continuous variables and the number with percentage was calculated for categorical variables.

**Results:** A total of 500 participants responded to the questionnaire. More than half of the responses were received from females (65%). On assessment it was found that, 26 % of respondents reported severe to extremely severe depressive symptoms; 31.5 % of respondents reported severe to extremely severe anxiety symptoms, and 19 % reported severe to extremely severe stress levels.

**Conclusion:** In India during the outbreak of COVID-19, an alarming number of students were found to have an impact on mental health due to the outbreak and were observed to have higher levels of stress, anxiety, and depression. The study findings shows the need of conducting more such studies and can be used to prepare appropriate psychological interventions to improvemental health among the young public during the pandemic.

## Introduction

In December 2019, a cluster of pneumonia cases of unknown cause was was reported to WHO which were identified in Wuhan city, China. [1] The organism responsible for this disease was discovered as a novel coronavirus (2019 n-CoV). Based on the genetic structure of the virus it was named as Corona (meaning crown) is a group of viruses.[2] This virus was found to cause respiratory symptoms and extra-pulmonary symptoms based on the type of the coronavirus. The virus has initially spread within few cities of China and later to many other countries, which made it a pandemic. [3] Epidemic such as Severe Acute Respiratory Syndrome Coronavirus (SARS-CoV) and the Middle East Respiratory Syndrome Coronavirus (MERS-CoV) which had occurred in previous years with other coronaviruses are considered as similar kind of disease. [2]

On 11^th^ February 2020, WHO declared the name of this disease as “COVID 19”;later the virus was renamed by the International Committee on Taxonomy of Viruses (ICTV) as Severe Acute Respiratory Syndrome Coronavirus 2 (SARS-CoV 2). Even though the novel corona virus is genetically related to the SARS-CoV outbreak of 2003 but it is of different type in several aspects. [4] The COVID-19 outbreak created a major panic among all the citizens of the country owing to its severity, contagiousness within the community, lack of specific treatment and possibility of re-infection. All these factors along with the uncertain behaviour of the virus lead to state of fear and concern all throught out the nation. The complete dynamics of transmission are yet to be determined but the general transmission of respiratory viruses happens through droplets.[3,5] Since the droplets travel approximately 1-meterin air and quickly settles on the surfaces, person to person transmission possibly occurs between close contacts. It is advised to frequently clean the hands with soap or alcohol-based sanitizers and to follow stand hygiene measures because the contaminated hands is reported to carry the virus into the body.[4, 5]

The known symptoms range from mild (cough, shortness of breath, etc) to severe (pneumonia, kidney failures, and death). Currently, there is no specific drug or vaccine,and the development of treatment and vaccines are under progress. [3] Since COVID-19 is a newly identified coronavirus, studies are going on to obtain more information about this organism as most of the data related to it is unknown. Hence all the above factors contribute to the agitation among the young generation in overall well being.

Currently, there is no much data on the mental health of the young public during the outbreak of COVID-19. This is especially relevant; uncertainty surrounding an outbreak of such a large extent. Mainly the research studies during the COVID-19 outbreak focuses on identifying the clinical features and epidemiology of infected patients [6], the genomic characterization of the virus [7], and challenges for worldwide health governance [8]. However, to our knowledge, published articles are examining the impact of COVID-19 on the young public of India.

Due to the widespread of the coronavirus in the public, the government of India has declared 21 days of lockdown in the first phase and 19 days of lockdown in the second phase for the entire country. As of lockdown, the people have to stay at home and during the lockdown, all the schools and colleges have been locked. The lockdown means isolation and to maintain social distancing to stop the spread of the infection. Sudden isolation and social distancing can significantly affect the mental health of people due to various reasons.

This study represents probably the first mental health survey conducted in the students of South Indiaafter the one month quarantine period of the COVID-19 outbreak. This study aims to find the prevalence of psychiatric symptoms among students. The results obtained may supportacademic institutions and healthcare professionals in safeguarding the psychological wellbeing of the studentsduring COVID-19 outbreak expansion in all parts of India.

## Methods

The present study is a cross-sectional, web-based online survey was conducted between 23rd April and 30th April 2020 (after one month period of lockdown). A 21-item DASS questionnaire was used to assess the emotional states of depression, anxiety, and stress. It is a set of three self-report scales; each of the three DASS-21 scales contains 7 items, divided into subscales with similar content.

A short form (21 items) and a long-form (42 items) which are valid and reliable measures inpatient and general population [9,10] different racial and cultural groups. This is a 21-item scale measured on a 4-point rating scale (0-3), “3” denoting “applied to me very much or most of the time”and“0”denoting“did not apply to me at all”. All the questions were provided with multiple options with only one answer to be chosen.

Using Google Forms, the questionnaire was randomly distributed among the pharmacy students of selected colleges. Through emails and WhatsApp, we requested people to circulate the survey link among their respective college mates. Participants were asked to give consent before taking part in the survey. The study ensured the confidentiality of the participant’s personal information they provided. The study does not deliver any intervention to participants; thus, there is no risk of physical harm to participating individuals. This study received an exemption from the Institutional Human Ethics committee. Mean with standard deviation was calculated for continuous variables and the number with percentage was calculated for categorical variables.

## Results

A total of 500 participants responded to the questionnaire. More than half of the responses were received from females (65%). The majority 85% of participants were from nuclear family and around 69 % are from the urban area. The age of participants ranged from 18 – 24 years; with mean (SD) age of 21.2 (+/- 1.3) years.

**Table 1:**
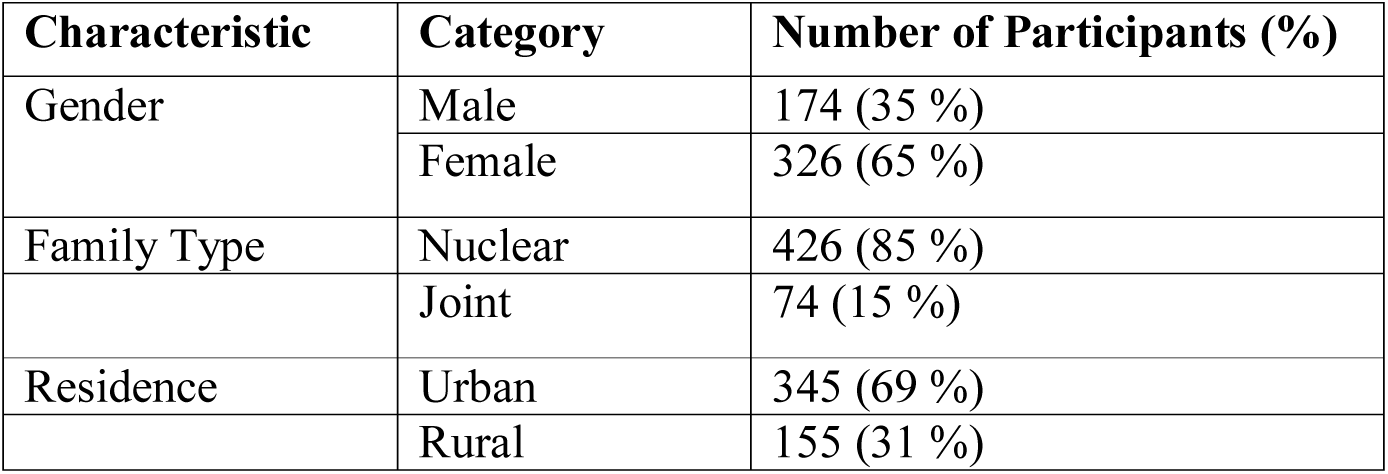
Demographic details of the study population

After the country’s outbreak of COVID-19, the government of India declared public health emergency of National concern, 26 % of respondents reported severe to extremely severe depressive symptoms; 31.5 % of respondents reported severe to extremely severe anxiety symptoms,and 19 % reported severe to extremely severe stress levels.

**Table 2:**
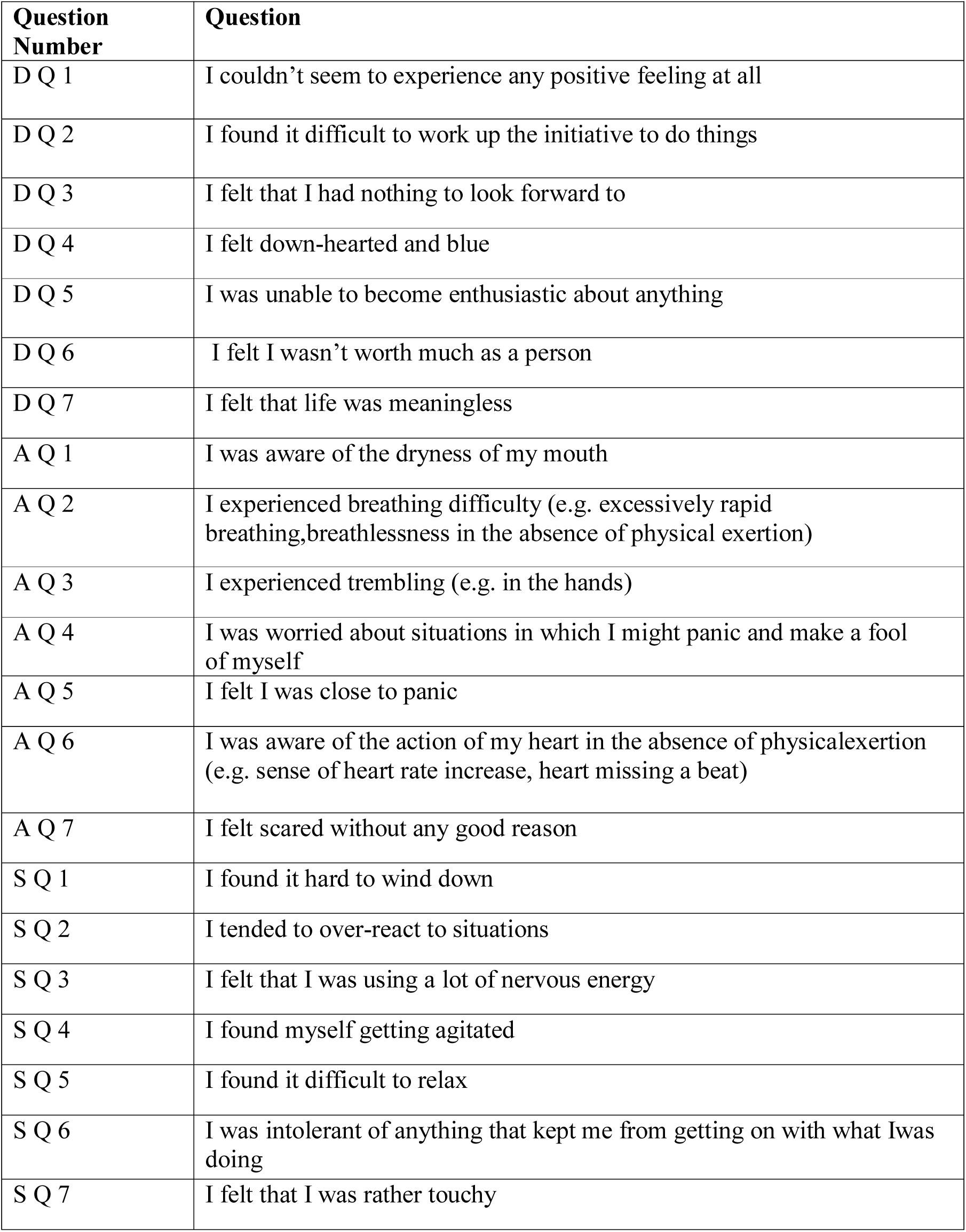
DASS-21; Set of questions (DQ: depression question, AQ: anxiety question, SQ: stress question), responses are rated from 0 to 3

The depression scale assesses the symptoms such as dysphoria, devaluation of life, hopelessness, self-deprecation, anhedonia, lack of interest/involvement and inertia. The anxiety scale assesses autonomic arousal, situational anxiety, skeletal muscle effects and subjective experience of anxious affect. The stress scale is sensitive to levels of chronic non-specific arousal. It assesses difficulty relaxing, nervous arousal and being easily upset/agitated, irritable / over-reactive, and impatient.

**Table 3:** Scores of the scale

|  | **Normal** | **Mild** | **Moderate** | **Severe** | **Extremely severe** |
| --- | --- | --- | --- | --- | --- |
| **Depression** | 0-9 | 10-13 | 14-20 | 21-27 | 28+ |
| No. Respondents (%) | 287 (57.5) | 42 (8.5) | 40 (8) | 41 (8) | 90 (18) |
| **Anxiety** | 0-7 | 8-9 | 10-14 | 15-19 | 20+ |
| No. Respondents (%) | 265 (53) | 25 (5) | 52 (10.5) | 20 (4) | 138 (27.5) |
| **Stress** | 0-14 | 15-18 | 19-25 | 26-33 | 34+ |
| No. Respondents (%) | 340 (68) | 20 (4) | 45 (9) | 33 (6.5) | 62 (12.5) |

## Discussion

Scores for all the 3 subsets of depression, anxiety, and stress are calculated by summing the scores obtained from the respondents for the relevant items. The DASS-21 is based on a dimensional rather than a categorical conception of psychological disorder. The assumption on which the DASS-21 development was based is that the differences between the depression, anxiety, and the stress experienced by normal subjects and clinical populations are essentially differences of degree which was confirmed by research data. This scale has no direct inferences for the distribution of patients to separate diagnostic categories proposed in classificatory systems such as the DSM and ICD.

Students were also found to havean impact on the outbreak and higher levels of stress, anxiety, and depression. As the total number of people infected by COVID-19 is rising in an alarming condition, major cities in South India have shut down all academic institutions at all levels indefinitely. The potential negative impact and uncertainity on academic development could hurt the mental health of students [11]. As young people are more receptive to digital applications [12], healthcare providers could consider providing online or smartphone-based psychological interventions and psychoeducation such as cognitive behavior therapy; to reduce the risk of negative impacts associated with the epidemic. A support network could be provided through online platforms for those people who are in quarantine during the epidemic. Health authorities need to provide needful information in a systematic format in simple languages (either audio or diagrammatic) to support the public with lower grades of educational background during the epidemic.

Though life needs stress constructively up to a certain limit and may be adequate for personality development, but if these stresses become too severe which disengage the psychic equilibrium producing maladaptive patterns of behavior. Inadequate interaction with the environment and family leads to stress and anxiety [13].

### This study has a few limitations

The resources available are limited in this period and we have adopted the snowball sampling strategy for collecting the data. As a result, huge data was not collected; the conclusion was less generalizable to the entire population, particularly less diverse young public. One more limitation of the study is that self-reported levels of anxiety, depression, and stress may not always be aligned with the assessment done by trained health professionals. The study also did not assess the risk factors which might have contributed for alteration of the mental health status.

This study provides important information about the respondents on the psychological responses of 4 weeks after the outbreak of COVID-19. Our study findings directly inform the need for the developing and implementing the strategies of several psychological interventions that may help minimizing anxiety, depression, and stress during the outbreak of COVID-19. As the COVID-19 epidemic, which is still ongoing at the time of communicating this manuscript, all the interventions developed will be helpful in large extent to Indian youth.

## Conclusion

In India during the outbreak of COVID-19, around 18 % of the respondents reported extremely severe depression, and about 27.5 % of respondents reported extremely severe anxietyand about 12.5 % of respondents reported extremely severe stress. An alarming number of students were found to havean impact on mental health due to the outbreak and were observed to havehigher levels of stress, anxiety, and depression. The study findings can be used to prepare psychological interventions to improvemental health among the young public during the COVID-19 epidemic.

## Data Availability

The authors confirm that the data supporting the findings of this study are available within the article

## Acknowledgment

The authors would like to thank all the study respondents for their active participation in providing the scientific data which helps in developing strategies/interventions for building a healthy Nation.

---

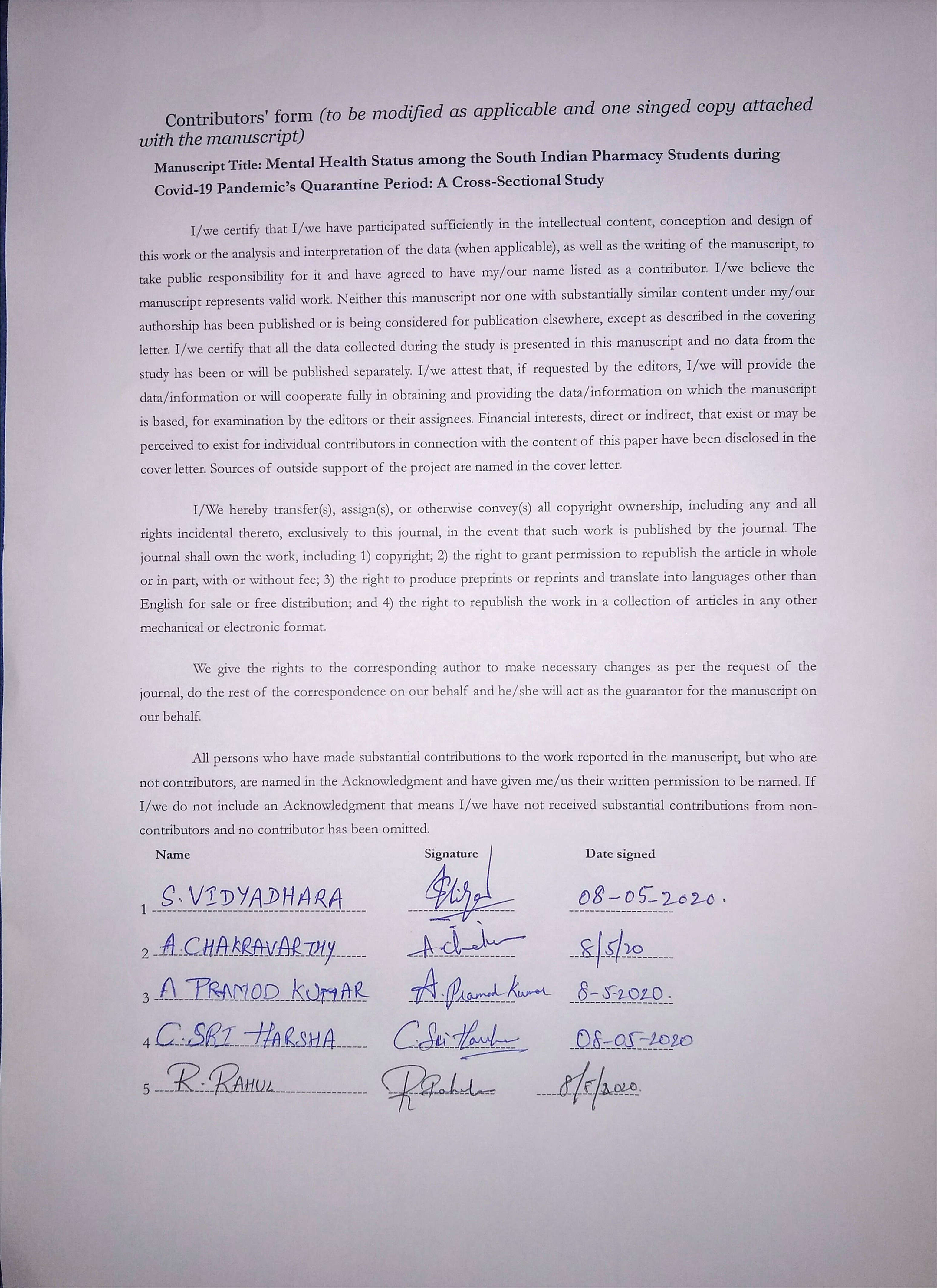

